# A Randomized, Single-blind, Group sequential, Active-controlled Study to evaluate the clinical efficacy and safety of α-Lipoic acid for critically ill patients with coronavirus disease 2019(COVID-19)

**DOI:** 10.1101/2020.04.15.20066266

**Authors:** Ming Zhong, Aijun Sun, Ting Xiao, Ge Yao, Ling Sang, Xia Zheng, Jinyan Zhang, Xuejuan Jin, Lei Xu, Wenlong Yang, Peng Wang, Kai Hu, Dingyu Zhang, Junbo Ge

## Abstract

**Object:** To evaluate the clinical efficacy and safety of α-Lipoic acid (ALA) for critically ill patients with coronavirus disease 2019 (COVID-19).

**Methods:** A randomized, single-blind, group sequential, active-controlled trial was performed at JinYinTan Hospital, Wuhan, China. Between February 2020 and March 2020, 17 patients with critically ill COVID-19 were enrolled in our study. Eligible patients were randomly assigned in a 1:1 ratio to receive either ALA (1200 mg/d, intravenous infusion) once daily plus standard care or standard care plus equal volume saline infusion (placebo) for 7 days. All patients were monitored within the 7 days therapy and followed up to day 30 after therapy. The primary outcome of this study was the Sequential Organ Failure Estimate (SOFA) score, and the secondary outcome was the all-cause mortality within 30 days.

**Result:** Nine patients were randomized to placebo group and 8 patients were randomized to ALA group. SOFA score was similar at baseline, increased from 4.3 to 6.0 in the placebo group and increased from 3.8 to 4.0 in the ALA group (P=0.36) after 7 days. The 30-day all-cause mortality tended to be lower in the ALA group (3/8, 37.5%) compared to that in the placebo group (7/9, 77.8%, P=0.09).

**Conclusion:** In our study, ALA use is associated with lower SOFA score increase and lower 30-day all-cause mortality as compared with the placebo group. Although the mortality rate was two-folds higher in placebo group than in ALA group, only borderline statistical difference was evidenced due to the limited patient number. Future studies with larger patient cohort are warranted to validate the role of ALA in critically ill patients with COVID-19.

## Introduction

Since December 2019, the novel coronavirus pneumonia 2019 (COVID-19) induced by severe acute respiratory coronavirus 2 (SARS-CoV-2) has become a worldwide pandemic and is overwhelming health care systems globally. Thus far, no therapeutics have yet been proven effective for the treatment of severe COVID-19, and the mortality rate of severe COVID-19 patients is high[1-5]. Pathological results suggested that the lung displayed diffuse alveolar damage. In addition, interstitial mononuclear inflammatory infiltrates, mainly lymphocytes, were observed in both lungs[6]. Moreover, several studies have shown that CRP, D-dimer and serum proinflammatory cytokines (IL-6, IL-1β, etc.) was increased in patients with severe COVID-19[6-9]. These findings suggested that the cytokine release syndrome (CRS) may be associated with COVID-19. CRS is a systemic inflammatory response that can be induced by infection and certain drugs. The sharp increase of pro-inflammatory cytokines is the main manifestation, which can cause damage to the heart, lungs and other organs[10]. Therefore, regulating systemic inflammatory response may be a key method for treating patients with COVID-19, and several clinical studies targeting CRS have been conducted[11]. Viral infection can also cause the production of reactive oxygen species (ROS), and ROS plays an important role in virus replication and invasion[12], organ damage[13, 14], and systemic inflammatory response[15-20].

α-Lipoic acid (ALA), as an antioxidant, has been confirmed to reduce systemic inflammatory response in patients with acute coronary syndromes, liver transplantation patients, and kidney-pancreas combined transplantation patients[21-24]. Therefore, the aim of this study was to investigate whether ALA could alleviate systemic inflammatory response by reducing ROS production, thereby improve organ function and the prognosis of critically ill patients with COVID-19.

## Methods

### Study design and participants

This study was a randomized, single-blind, group sequential, active-controlled trial. Participants were recruited from February 2020 to March 2020. 17 participants with critically ill COVID-19 hospitalized in JinYinTan Hospital, Wuhan, China were enrolled. Ethical approval was obtained from the Institutional Ethics Committee of Zhongshan Hospital(B2020-030). The informed consent was signed by all participants. The trial has been registered in the Chinese Clinical Trial Registry (ChiCTR2000029851). The inclusion criteria were: 1. Patients diagnosed with critically ill COVID-19. It complies with the COVID-19 Critical and Critical Diagnostic Standards, namely “Pneumonitis Diagnosis and Treatment Scheme for Novel Coronavirus Infection (Trial Version 5); 3. Sign written informed consent. Patients who cannot sign informed consent must obtain informed consent from the independent authorized nurse. The exclusion criteria were: 1. Patients who are participating in other clinical trials; 2. Pregnant or breastfeeding women; 3. There are other life-threatening diseases such as cancer; 4. Expected survival time <24 hours; 5. Patients who are allergic to ALA or similar drugs (B vitamins), and intolerant to the recommended dosage of ALA in the past; 6. A history of immune system diseases or diseases closely related to the immune system (Figure 1).

**Figure. 1.**
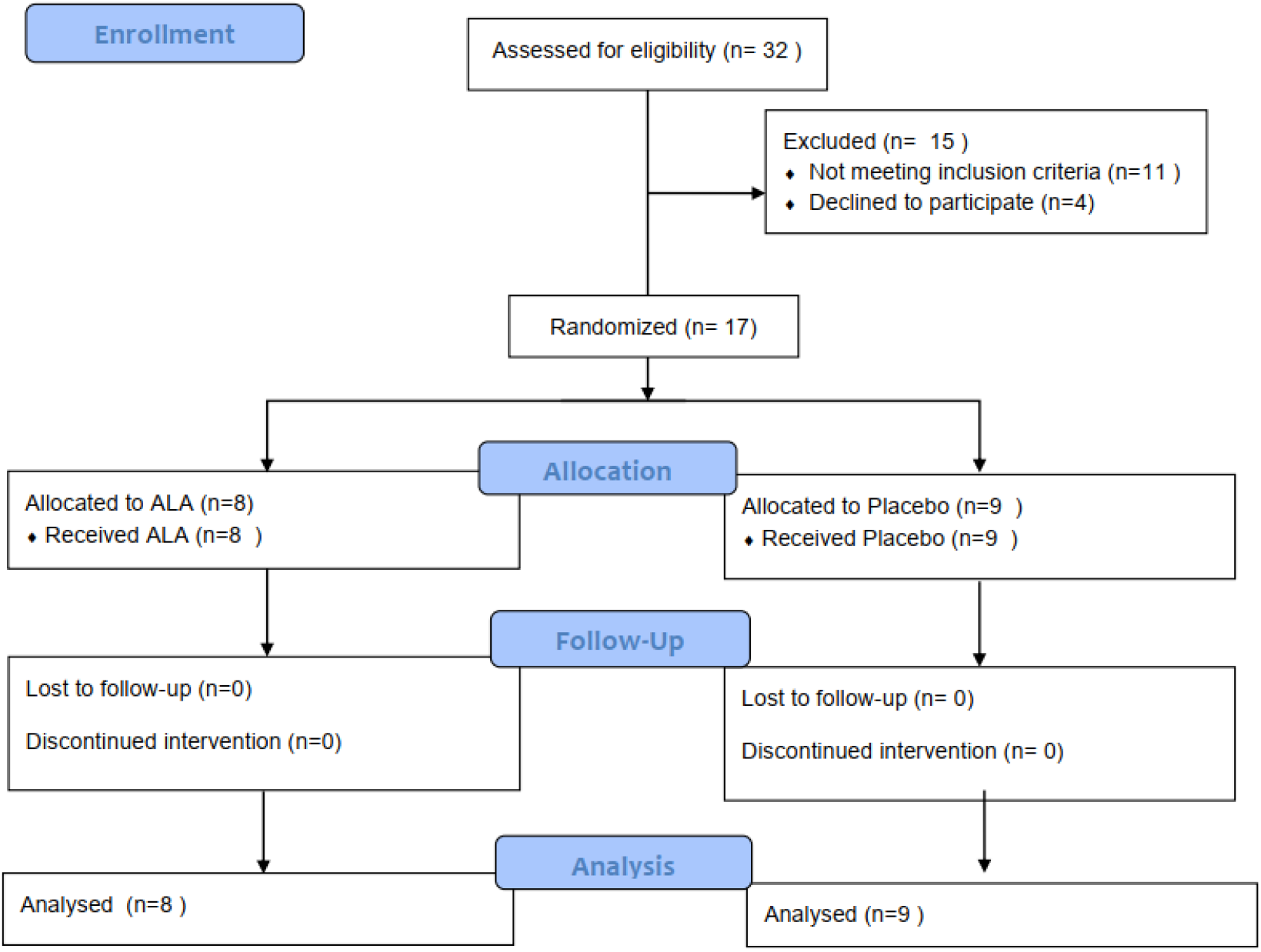
Flow Diagram

### Randomization and Blinding

We used mixed randomization for unpredictability in allocation sequences[25]. The allocation sequences were placed in the sealed opaque envelopes. We randomly assigned critically ill patients with COVID-19 to the treatment group and control group in a 1:1 ratio. Blinding is saved until research have been completed and the database is finally unlocked. Participants and study personnel were aware of the study-group assignments, but outcome adjudicators were not.

### Procedures

Patients in the treatment group were treated with ALA (1200 mg/d, intravenous infusion) once daily plus for 7 days on top of standard medical care. Patients in the control group were treated with equal volume saline infusion on top of standard medical care for 7 days. ALA is discontinued when the treatment plan is completed or the patient meets the halfway withdrawal criteria. Withdrawal criteria: (1) The subject himself requested to withdraw from the trial; (2) An intolerable adverse event or an adverse event that the investigator believes must be withdrawn from the study; (3) Those who cannot be treated according to the protocol, have poor compliance, or the physician believes that they should withdraw from the trial.

### Clinical and Laboratory Monitoring

Initial investigations included onset time, CT results, nucleic acid test results, previous history, heart-related symptoms (chest tightness, etc.), NYHA classification, personal history, comorbidities and treatment plans, comprehensive physical examination (including blood pressure, heart rate, weight, oxygen Saturation measurement), Sequential Organ Failure Estimate (SOFA) Score, electrocardiogram results, echocardiogram results and laboratory test results (blood routine, liver and kidney function, electrolytes, blood glucose, blood lipids, CRP, urine pregnancy test (women), brain natriuretic peptide, myocardial enzymes, troponin, coagulation function, serum inflammatory factor tests). During the clinical follow-up period (days 1, 2, 3, 4, 5, 6, 7), the following data were collected from patients each day: symptoms, blood pressure, heart rate, fingertip oxygen saturation (recorded twice daily: 9am and 3pm), SOFA score, major and minor endpoint event records, adverse event records and laboratory tests (blood routine, Liver and kidney function electrolytes, myocardial enzymes, troponin, NT-proBNP, CPR, coagulation).

### Primary outcomes and Secondary outcomes

The primary outcome of this study was the SOFA Score, and the secondary outcome was the all-cause mortality up to 30 days. All outcomes were determined by an independent clinical endpoint determination committee.

### Statistical analysis

We estimated that with 10 deaths the study would have 68% power to detect a 50% improvement in survival with ALA versus standard control group (from admission to discharge), at a two-sided alpha level of 0.05. Primary and secondary endpoints were measured in the intention-to-treat population, which included all patients who underwent randomization. The Kaplan–Meier method was used to estimate time-to-event end points. Two-sided stratified log-rank tests and 2Log (LR) test were used. Cox regression models were applied to estimate hazard ratios. Safety evaluations were included all patients who received at least one dose of study medication. Continuous variables were described using mean, median, and standard deviation, and examined using Analysis of Variance (ANOVA) or Wilcoxon rank sum test or Kruskal-Wallis test. Categorical variables were described by frequency, and assessed using chi-square test or Fisher exact test. Changes in laboratory data, signs and examination items, and adverse events before and after treatment in the two groups will be analyzed as safety indicators. This study does not estimate missing data. Statistical analysis was performed using SAS 9.13. Values of *P* < .05 were considered statistically significant, and the two-sided test is represented by a 95% confidence interval.

## Results

### Characteristics of the Patients

A total of 17 eligible patients with critically ill COVID-19 were enrolled in our study. 8 patients were assigned to receive ALA and 9 patients to standard care alone. A total of 13 patients (76.5%) were men, the age range was 51 to 91 years, and the median age of patients was 63 years (interquartile range [IQR], 59 to 66 years) (Table 1). There were no significant differences between groups in demographic characteristics, baseline laboratory test results and SOFA Score at enrollment.

**Table 1:**
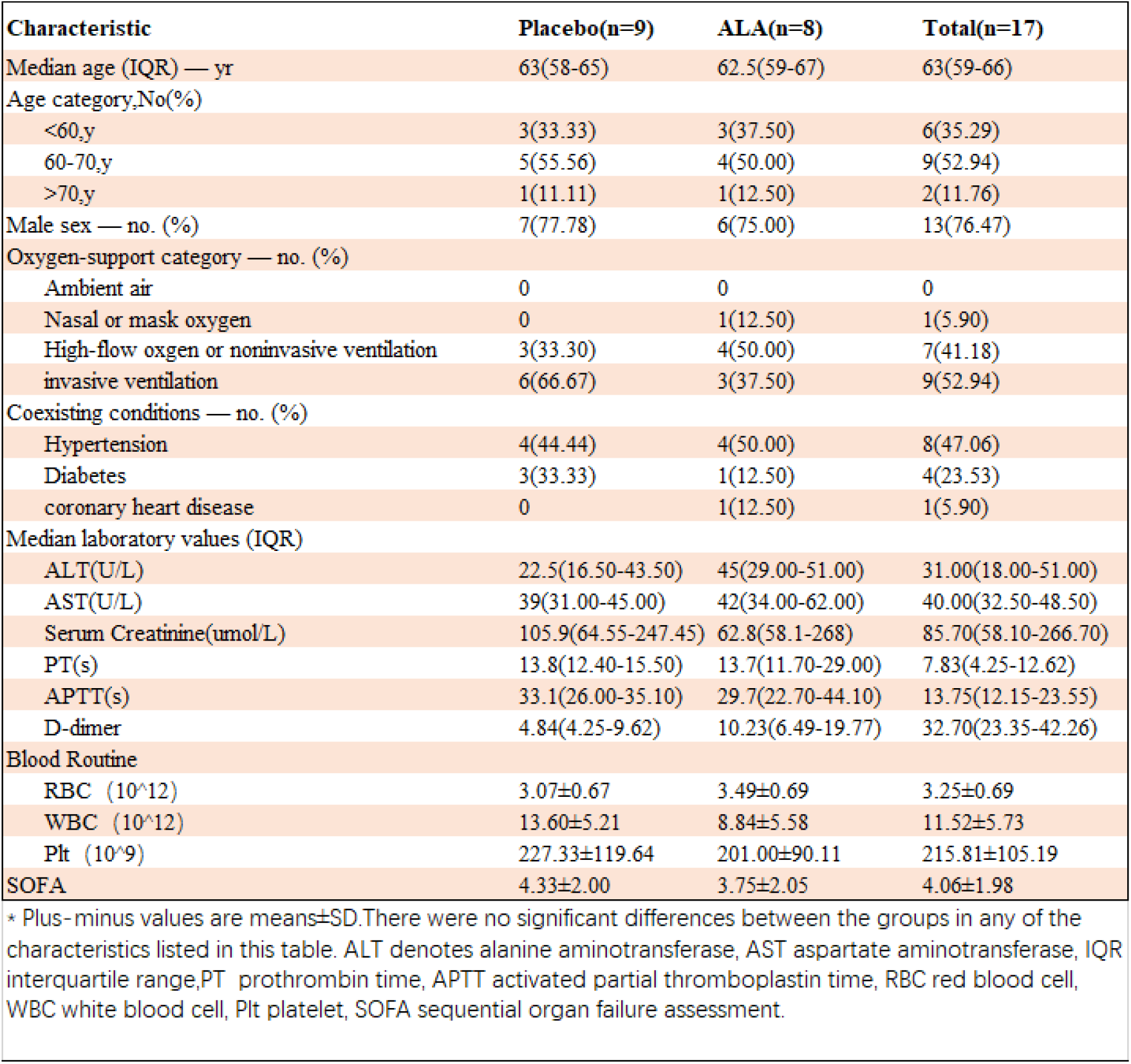
Demographic and Clinical Characteristics of the Patients at Baseline*

### Primary Outcomes

From day 0 to day 7, the average SOFA Score in the placebo group increased by 1.7 points from 4.3 to 6.0. In the same time period, the average SOFA Score in the ALA treatment group increased by 0.2 points from 3.8 to 4.0 (Figure 2). There was no significant difference in SOFA Score between the placebo group and the ALA group (p=0.36).

**Figure. 2.**
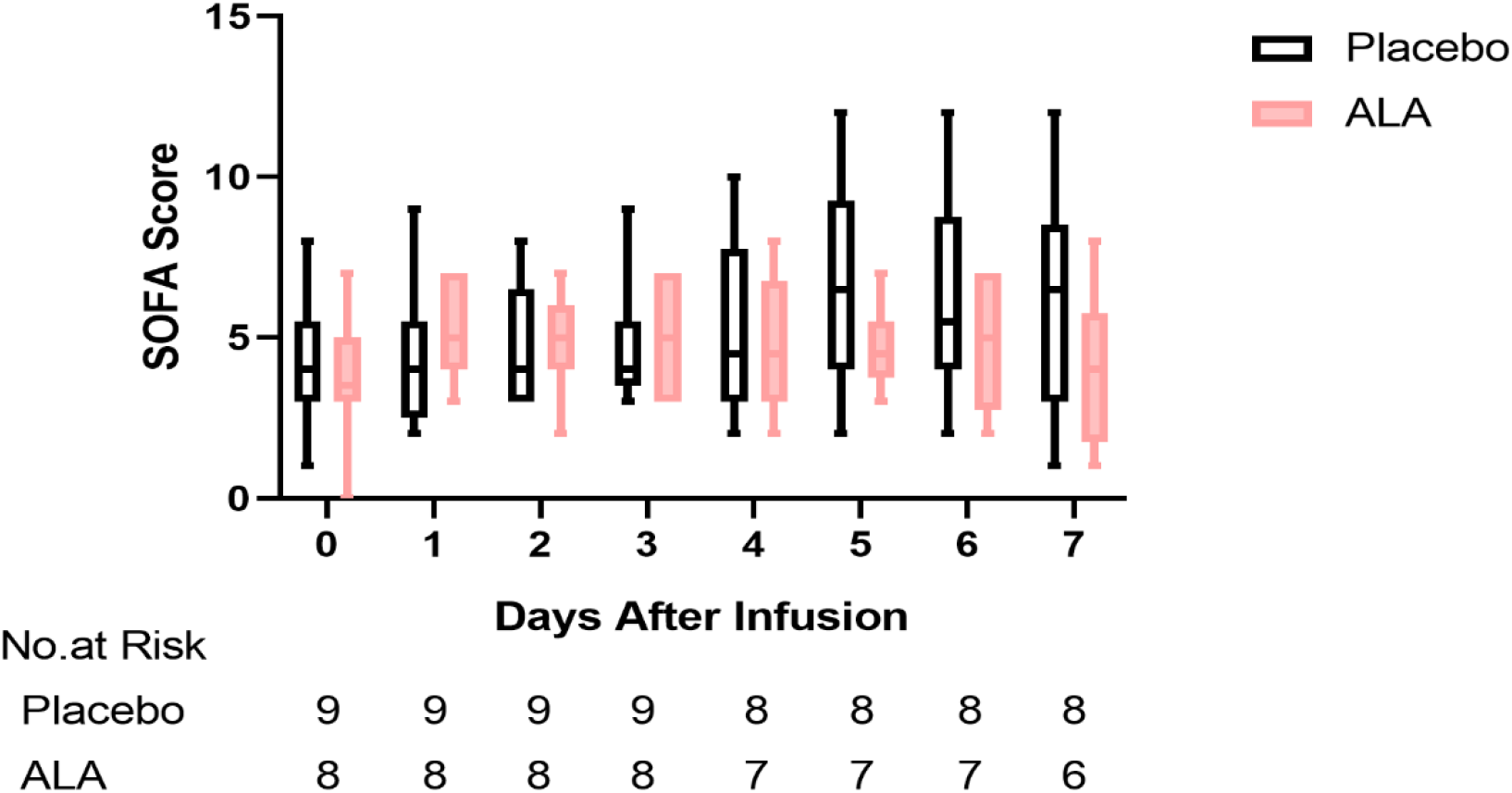
SOFA score

### Secondary Outcomes

After a 30-day follow-up, the 30-day all-cause mortality was 77.8% (7/9) in the placebo group, and 37.5% (3/8) in the ALA group (Figure. 3). The 30-day all-cause mortality was numerically lower in the ALA group than in the placebo group, a borderline statistical significance was observed (P=0.09).

**Figure. 3.**
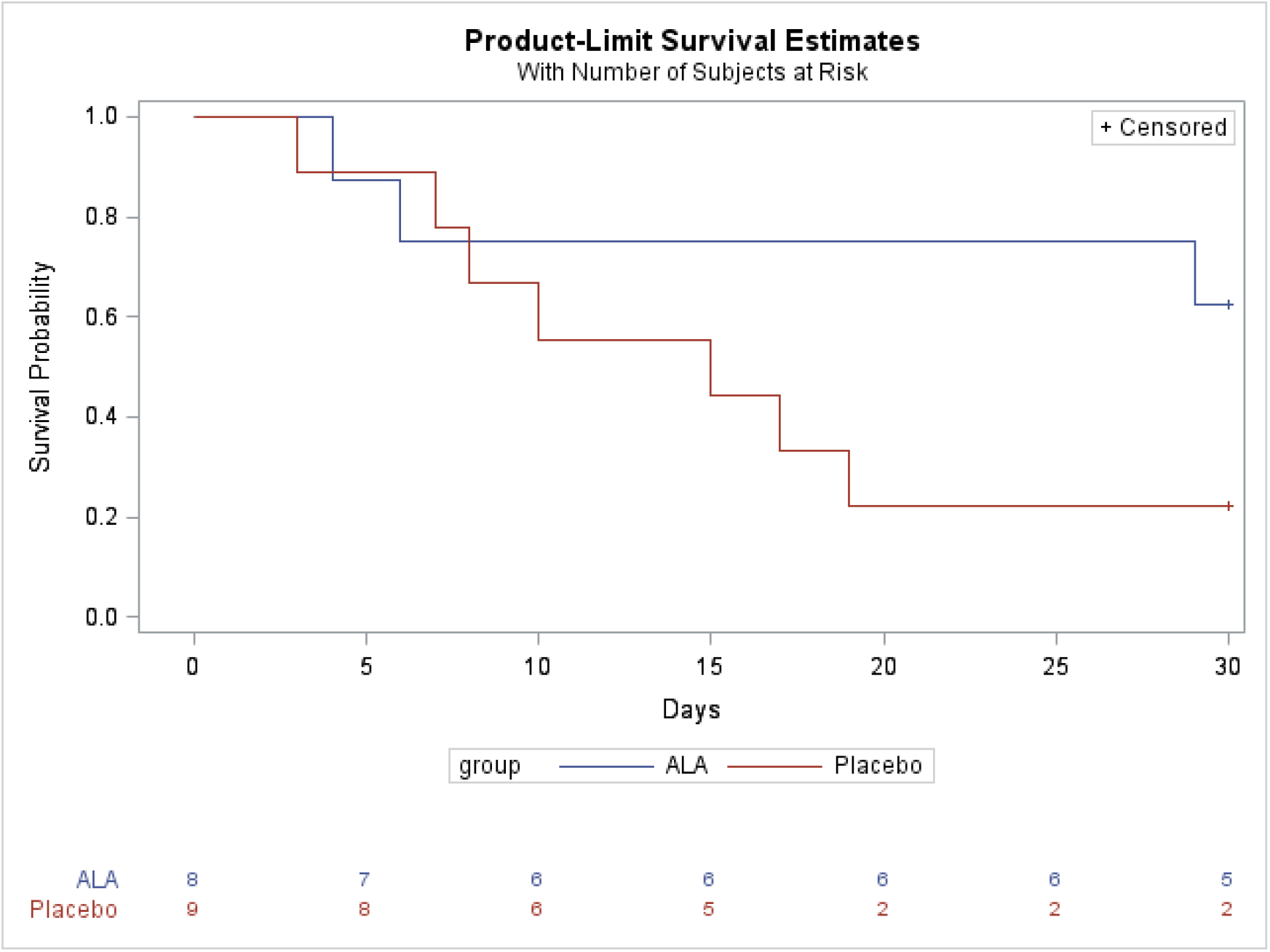
Overall Survival; Borderline significance was observed between ALA group and placebo group (P=0.09) at day 30.

### Adverse Events

Patients were followed up for adverse events. No unexpected drugs-related adverse events occurred during the trial.

## Discussion

Our study enrolled 17 patients with critically ill COVID-19 in Wuhan JinYinTan Hospital. The results suggested ALA treatment might be able to improve 30-day survival rate of patients with critically ill COVID-19 and slow down the increase of SOFA score, both parameters did not reach statistical significance due to the limited patient number, but the tendency of improvement is clear to see.

The protection efficiency of ALA in critically ill COVID-19 patients might be explained in two aspects, antioxidant and anti-inflammation. Firstly, ALA could ameliorate virus-induced organ dysfunction by counteracting ROS. Ross[14] revealed that excessive superoxide induced by influenza A virus infection could cause organ injury, which could be alleviated by inhibiting ROS production. Besides, Xian and colleagues[12] suggested that the replication of H5N1 influenza virus was impeded by overexpression of Cu/Zn superoxide dismutase (SOD1), which was one of the antioxidation enzymes. Secondly, the anti-inflammation effect of ALA was verified by a series of clinical trials. ALA could significantly reduce levels of serum inflammatory cytokines and improve symptoms of some severe patients (such as acute coronary syndrome, etc.)[21-24]. Some clinical trials have been designed to dampen inflammatory responses. One clinical trial (ChiCTR2000029765) reported that neutralizing IL-6 with tocilizumab could quick control of high fever and respiratory symptoms in 21 patients with COVID-19[11], which revealed feasibility of anti-inflammation treatment for COVID-19.

The clinical trials of hydroxychloroquine combined with azithromycin in the treatment of COVID-19 by Molina et al[29] (N = 11) focused on patients with mild COVID-19. In another clinical trial focused on patients with severe COVID-19, there was no significant difference in viral RNA loads or duration of viral RNA detectability between the lopinavir-ritonavir combination treatment group and the control group, but lopinavir-ritonavir treatment group presented with lower all-cause mortality on day 28 (P>0.05)[30]. Thus, in addition to the anti-virus treatment strategy, searching for other treatment strategies are of clinical importance aiming to alleviate the complications of COVID patients and reduce the mortality rate of patients with severe and critically ill COVID-19 was also essential.

This trial has several limitations. In particular, the trial did not have sufficient number of cases. Because the epidemic situation in Wuhan was rapidly under control during the trial period, and critically ill COVID patients participating other clinical trials could not be enrolled in this study, only 17 critically ill COVID patients were enrolled in this study. Another limitation is that we use all-cause mortality as our secondary outcome, which could not distinguish death due to primary SARS-COV-2 insult from secondary bacterial infection, which was common in late phrase of critically ill patients with COVID-19. Further clinical studies are needed to verify the promising efficacy of ALA injection in patients with critically ill COVID-19.

In conclusion, our results derived from the small number of critically ill COVID patients are promising and hope this report might stimulate the initiation of further clinical studies in larger patient cohort to validate the role of ALA on reducing the short-term mortality rate among critically ill COVID-19 patients.

## Data Availability

all data referred to in the manuscript was available

## Acknowledgement

We thank Mr. Huirong Zhang from YAOPHARMA CO., LTD, for the donation of α-Lipoic acid used in this study.

## Author Approval

This manuscript has been seen and approved by all listed authors.

## Data Availability Statement

all data referred to in the manuscript was available.

